# Precision psychological markers enable targeted treatment in digital eating disorder interventions

**DOI:** 10.64898/2026.07.21.26358594

**Authors:** Eric Hurwitz, Lindsay Merchant, Rachael E. Flatt, Kylie K. Reed, Zachary Butzin-Dozier, Melissa A. Haendel, Laura M. Thornton, Cynthia M. Bulik, Rachel Presskreischer

## Abstract

Digital mental health interventions for eating disorders can expand treatment access, but high attrition rates and heterogenous patient responses indicate a need for precision medicine approaches that identify which patients will benefit most and predict their treatment outcomes. We analyzed 30 days of Recovery Record (a widely-adopted digital intervention for eating disorders) data from 1,166 participants with lifetime bulimia nervosa or binge-eating disorder, with assessments at baseline, midpoint, and endpoint. We identified three symptoms (eating preoccupation, shape/weight preoccupation, and fear of losing control over eating) as precision psychological markers that predict and inform treatment response. These symptoms demonstrated the strongest correlations with overall symptom improvement measured by the Eating Disorder Examination Questionnaire (EDE-Q) Global score (*r*=0.65-0.66), predicted outcomes when elevated at baseline (all *P*<0.001), mediated treatment effects through early symptom changes (all *P*<0.001), and differentiated response groups in cluster analysis. Participants demonstrated significant improvements across all eating disorder domains at the population level (Global score Cohen’s *d*=-0.80), but cluster analysis revealed three distinct response patterns: strong responders (35.2%) achieved mean Global score reductions of -1.50 points, moderate responders (46.4%) achieved -0.36 points, and non-responders (18.4%) showed minimal change (-0.06 points). This framework may enable identification of individuals most likely to benefit from a digital intervention and potentially supports early treatment monitoring, paralleling precision medicine advances where psychological marker-driven patient selection could improve treatment decisions. This mechanistic approach provides a generalizable framework for developing targeted digital mental health interventions across psychiatric disorders.

## Introduction

Eating disorders are serious psychiatric conditions that carry significant morbidity and mortality^1^. Two of these disorders, bulimia nervosa (BN) and binge-eating disorder (BED) include episodes of binge eating, defined as consuming more than one would typically eat in a discrete period of time (e.g., 2 hours), and a sense of loss of control over eating^2,3^. BN also includes “compensatory behaviors” to prevent weight gain such as fasting, vomiting, laxative misuse, and excessive exercise^4^. Interventions such as cognitive-behavioral therapy (CBT) are effective for many people with BN and BED, and there is an FDA medication approved for the treatment of each (fluoxetine for BN, and lisdexamfetamine for BED)^5,6^. However, many people do not have access to these interventions due to geographic and financial barriers, stigma associated with seeking treatment, and provider shortages^7^. Strikingly, clinical trial outcomes for BN and BED are no better today than 40 years ago^8^.

As digital technologies spread across the globe, the health sector has sought ways to harness technology to expand access to interventions. These interventions have been designed for many types of media including CD-ROM, web-based, and mobile applications (apps)^9^. Lately, a large number focused on mobile apps as smartphones became ubiquitous. In a systematic review of treatment, self-monitoring, and multipurpose mental health apps, mixed results were found for their clinical effectiveness across a range of diagnoses, with attrition and non-completion (likely different across demographics and disease conditions) cited as key reasons for lower impact^10^. For digital eating disorder treatment interventions specifically, a systematic review noted that across all studies there was an approximate 25% attrition rate. Of three studies that reported on participant adherence, an average of 36% completed all study components^11^.

Despite the proliferation of digital mental health interventions, most adopt a “one-size-fits-all” approach that assumes equal benefit across all users. This contrasts sharply with advances in precision medicine in other medical fields. For example, in oncology, biomarker testing guides treatment selection, such as HER2 status determining whether breast cancer patients receive trastuzumab (Herceptin), a targeted therapy for HER2-positive tumors.^12^ This precision approach has transformed cancer care by matching treatments to patients most likely to benefit, improving both clinical outcomes and resource allocation.

Digital mental health interventions lack equivalent precision medicine frameworks for targeted treatment. Although thousands of mental health apps now exist, few are backed by rigorous research, and even fewer identify which users will benefit most^13^. This absence of personalization may partially explain the high attrition rates and modest effect sizes observed across digital intervention trials. Without prospective psychological markers to guide patient selection or early indicators to monitor treatment trajectory, clinicians cannot determine which individuals should be directed toward a specific digital intervention versus alternative care pathways. Users (and developers of these interventions) may invest time and effort in interventions unlikely to see benefits, while healthcare systems struggle to allocate resources efficiently and effectively.

Recovery Record is a mobile application used by over 2M people designed to support eating disorder recovery through CBT principles, including meal logging, emotion tracking, behavior monitoring, and real-time coping skill delivery^14,15^. Previous research has demonstrated Recovery Record’s effectiveness, with approximately 60% of users achieving clinically meaningful improvement in eating disorder symptoms^16^. However, these aggregate efficacy estimates obscure heterogeneity in treatment response. Some individuals experience dramatic symptom reduction while others show minimal change, yet no framework exists to prospectively identify who will benefit most or to understand the mechanisms driving differential response.

This study addresses the precision medicine gap in digital eating disorder interventions by developing a mechanistic framework to identify clinical markers (i.e., specific symptoms) predicting Recovery Record response. We hypothesized that there would exist specific symptoms that function as precision psychological markers (analogous to HER2 in breast cancer) that are predictive of individuals responding to the Recovery Record intervention and can be used to elucidate the intervention’s therapeutic mechanisms. By establishing which baseline symptom profiles predict response and which early symptom changes drive sustained improvement, we aimed to create a framework enabling evidence-based treatment matching and early trajectory monitoring. Such a framework could transform digital eating disorder care from reactive (waiting for treatment failure) to proactive (early identification and redirection of unlikely responders), paralleling precision medicine advances in other medical fields. Beyond eating disorders, this systematic approach to psychological marker identification may provide a generalizable template for developing precision frameworks across digital mental health interventions.

## Methods

### Study data and participants

Data in this study were analyzed as part of the Binge Eating Genetics Initiative (BEGIN) study (approved by University of North Carolina Biomedical Institutional Review Board Protocol #17-0242)^17^. Participants were recruited nationally via Recovery Record, social media, the National Eating Disorders Association, and a University of North Carolina participant registry listserv from August 2017 to March 2021^17,18^. Eligibility was determined by the ED100k.v2 and required a lifetime diagnosis of BN or BED based on DSM-5 criteria, current binge-eating behavior, age 18–45 years, and access to an iPhone^17,19^. Enrolled participants were mailed study materials including an Apple Watch device (1st generation); participants were permitted to keep the device provided to them upon study completion and were not otherwise compensated^17,18^. The study involved a 30-day intervention period with assessments at three timepoints: baseline (Timepoint 1, Day 0), midpoint (Timepoint 2, Day 15), and endpoint (Timepoint 3, Day 30). The final analytic sample comprised 1,166 participants who contributed complete baseline data.

Notably, participants were recruited in part through Recovery Record itself, which pushed study notifications to existing users, and were required to complete three logs in the app before being eligible to enroll. Consequently, a large subset of participants likely had prior app familiarity at study entry^18^. No data were collected on prior Recovery Record use, and differential prior app exposure should be considered when interpreting baseline engagement and treatment response.

### Intervention

Recovery Record is a mobile platform supporting eating disorder recovery through core CBT principles, enabling users to track meals, eating disorder behaviors, thoughts and feelings, and progress toward goals, and providing access to coping skills, affirmations, and reminders^14^. Participants had unrestricted access to all app features following enrollment. Data collection was limited to the 30-day intervention period, though participants were free to continue using the app thereafter.

### Demographic and clinical variables

Age was categorized into five-year bins (18-24, 25-29, 30-34, 35-39, 40-45 years), consistent with CDC age groupings used in national health surveillance; the wider terminal bins reflect study eligibility boundaries^20^. Sex was determined by genotype when available or by self-report^21^. Race and ethnicity were combined into a single variable consistent with the CDC; participants identifying as Hispanic or Latinx were categorized as Hispanic/Latinx of any race, with all remaining participants classified by race: White non-Hispanic (NH), Black NH, Asian NH, Native American NH, more than one race NH, and Unknown NH^22^. Additional clinical variables collected (from the ED100k.v2) at baseline included lifetime eating disorder diagnoses, treatment history, medication use, compensatory behavior frequency, anthropometric data (current body mass index [BMI], highest lifetime BMI, lowest adult BMI), and family history of eating disorders^17^.

### Outcome measures

The primary outcome was eating disorder psychopathology assessed using the Eating Disorder Examination Questionnaire (EDE-Q), a validated 28-item self-report measure generating a Global score (0–6) representing overall symptom severity and four subscale scores: Restraint, Eating Concern, Shape Concern, and Weight Concern^23,24^. Secondary outcomes were depression symptoms assessed by the Patient Health Questionnaire-9 (PHQ-9; 0–27) and anxiety symptoms assessed by the Generalized Anxiety Disorder-7 (GAD-7; 0–21), both with well-established validity and reliability^25,26^. Higher scores on all measures indicate greater symptom severity. Detailed item content for the EDE-Q, PHQ-9, and GAD-7 is provided in Tables S1 and S2.

### Statistical analysis

To examine symptom changes over time, we fit linear mixed-effects models with random intercepts for each participant to account for within-person correlation across the three assessment timepoints^27^. Models were fit separately for each outcome measure. Post-hoc pairwise comparisons between all three timepoints were conducted using estimated marginal means. Cohen’s *d* effect sizes for within-person changes were calculated as the mean of within-person differences divided by their standard deviation and interpreted as negligible (|*d*| < 0.2), small (0.2 ≤ |*d*| < 0.5), medium (0.5 ≤ |*d*| < 0.8), and large (|*d*| ≥ 0.8)^28,29^. Item-level effect sizes were calculated for each EDE-Q, PHQ-9, and GAD-7 item across all three intervals (Timepoint 1-2, Timepoint 2-3, Timepoint 1-3).

App engagement was measured as the cumulative number of user-initiated interactions with Recovery Record during the 30-day period, extracted from timestamped event logs. We examined associations between engagement and outcome change scores (Timepoint 1 to Timepoint 3) using linear regression, adjusting for baseline age, sex, and race/ethnicity.

All analyses were conducted in R version 4.4.0^30^. Statistical significance was set at α=0.05, with Bonferroni correction applied to pairwise comparisons^31^.

### Precision psychological marker identification

We employed a four-pronged analytical framework to identify eating disorder symptoms serving as precision psychological markers for treatment response, designed to establish qualitatively distinct forms of evidence: 1) *correlational relevance* — symptom change correlates with overall EDE-Q Global score improvement; 2) *prospective prediction* — baseline symptom levels identify individuals most likely to respond before treatment begins; 3) *mechanistic mediation* — early symptom improvement (Timepoint 1-2) predicts continued EDE-Q Global score improvement in the later phase (Timepoint 2-3), establishing temporal precedence consistent with a causal role; and 4) *cluster differentiation* — the symptom distinguishes naturally occurring response subgroups. Symptoms meeting criteria across all four approaches were designated precision psychological markers.

#### Correlation Analysis

We calculated Pearson correlations between changes in individual EDE-Q items (Timepoint 1-3) and changes in EDE-Q Global scores. Based on item-level effect size analyses, we focused on six symptoms showing moderate sample-level improvements: eating preoccupation (Q7), shape/weight preoccupation (Q8), fear of losing control over eating (Q9), guilt about eating (Q20), weight importance (Q22), and shape importance (Q23) (Table S1), plus one symptom with negligible sample-level change as a negative control: reaction to prescribed weighing (Q24). The negative control tested specificity; uniformly strong correlations regardless of population-level effect would suggest non-specific associations rather than precision targeting.

#### Baseline Prediction

We stratified participants into high versus low baseline groups using median splits for each of the six candidate symptoms plus the negative control, then compared EDE-Q Global score change (Timepoint 1-3) between groups using independent-samples t-tests with Bonferroni correction. Symptoms showing significantly greater treatment response in the high-baseline group were considered candidate precision psychological markers.

#### Temporal Mediation

For each symptom, we calculated early symptom change (Timepoint 1-2) and late EDE-Q Global score change (Timepoint 2-3), then fit linear regression models predicting late Global change from early symptom-specific change, controlling for baseline EDE-Q Global severity. Significant negative β coefficients indicate that early symptom improvement drives subsequent EDE-Q Global improvement, supporting a causal mediation role. Statistical significance was assessed using Bonferroni correction.

#### Cluster Analysis

We performed hierarchical clustering using Euclidean distance on individual change trajectories for EDE-Q Global score and the three symptoms showing the strongest correlations with EDE-Q Global change (Q7, Q8, Q9)^32^. We evaluated solutions from k=2 to k=10 using the elbow method, silhouette analysis, and the gap statistic^33–35^. The elbow method identified k=3 as optimal; silhouette analysis yielded an average width of 0.26, indicating weak-to-moderate separation consistent with a continuum of response rather than discrete categories. We characterized the three clusters by mean change for EDE-Q Global scores and each of the three precision marker symptoms, and compared baseline symptom levels across clusters using one-way ANOVA with Bonferroni-corrected post-hoc tests. Baseline levels of the negative control symptom (Q24) were also compared to test specificity.

### Cluster characterization

To determine whether response clusters reflected distinct demographic or clinical subgroups, we characterized clusters across demographic and clinical variables described above captured from the ED100k.v2. Continuous variables were compared across clusters using one-way ANOVA with pairwise t-tests for variables showing significant overall effects. Categorical variables were examined using chi-square tests, with pairwise comparisons conducted for variables showing significant overall associations^36^. Bonferroni correction was applied for pairwise comparisons.

### Use of generative AI

We used ChatGPT (OpenAI) and Claude (Anthropic) to assist with language refinement and R code generation and debugging, in accordance with UNC guidelines^37–39^. No study data were submitted to these tools. All outputs were critically reviewed by the authors. These tools were not used for generating scientific concepts or ideas.

## Results

### Cohort characteristics

Our cohort included 1,166 individuals enrolled in the BEGIN study via the Recovery Record application. The cohort was predominantly female (86.5%), White Non-Hispanic (78.1%), and 18–24 years old (31.2%) (Table 1). Of the 1,166 participants, 740 (63.5%) logged at least one eating disorder-related behavioral event over the study period, recording 22,075 events in total (median 20 events per person, IQR=10-38) (Table 2). Meal skipping was the most frequently logged behavior (46.6% of events), followed by dietary restriction (26.5%), binge eating (23.7%), purging (2.7%), and laxative use (0.6%).

**Table 1:** Descriptive statistics of the cohort.

| **Group** |  | **Count (%)** |
| --- | --- | --- |
| **Total** |  | **1166 (100%)** |
| **Sex:** | Female | 1009 (86.5%) |
|  | Male | 154 (13.2%) |
| **Age:** | 18-24 | 364 (31.2%) |
|  | 25-29 | 258 (22.1%) |
|  | 30-34 | 221 (19.0%) |
|  | 35-39 | 187 (16.0%) |
|  | 40-45 | 136 (11.7%) |
| **Race/ethnicity:** | Black NH | 44 (3.8%) |
|  | Asian NH | 44 (3.8%) |
|  | White NH | 911 (78.1%) |
|  | More than one race NH | 129 (11.1%) |
|  | Native American NH | 34 (2.9%) |

Note, specific groups with counts <20 were omitted for privacy.

**Table 2:** Descriptive statistics of eating disorder events across the study period.

| Event | Number of individuals | Total number of events (%) | Median events per person (IQR) |
| --- | --- | --- | --- |
| **All events** | **740** | **22,075 (100%)** | **20 (10-38)** |
| Meal skipped | 645 | 10,284 (46.6%) | 10 (4-22) |
| Restricted | 500 | 5,850 (26.5%) | 6 (2-14) |
| Binge ate | 658 | 5,222 (23.7%) | 6 (3-10) |
| Self-induced vomiting | 85 | 595 (2.7%) | 4 (2-9) |
| Laxative misuse | 29 | 124 (0.56%) | 2 (1-3) |

### Individuals using Recovery Record experience improved eating disorder outcomes

Participants demonstrated significant improvements across all eating disorder domains over 30 days of Recovery Record use. EDE-Q Global scores decreased from 3.91 (95% CI [3.84, 3.97]) at baseline to 3.19 (95% CI [3.12, 3.26]) at endpoint (Cohen’s *d*=−0.80), with medium-to-large improvements across all subscales (Eating Concern: *d*=−0.80; Restraint: *d*=−0.59; Weight Concern: *d*=−0.55; Shape Concern: *d*=−0.51) (Figure S1). Modest improvements were observed for depressive symptoms (PHQ-9: *d*=−0.35), while anxiety showed minimal change (GAD-7: *d*=−0.09). Greater user engagement was associated with improved EDE-Q Global scores (β=-0.27, standard error [SE]=0.13, *P*=0.04) and EDE-Q Shape scores (β=-0.46, SE=0.16, *P*=0.004) (Figure S2). However, understanding *who* benefits and *which symptoms* drive that benefit requires moving beyond sample-level estimates, which is the focus of subsequent analyses.

### Eating disorder outcomes by specific symptom

To better understand the mechanisms underlying the broad improvements in eating disorder psychopathology among Recovery Record users, we examined changes in individual EDE-Q items and comorbid symptom domains. The top five symptoms (captured by individual questions) with the largest effect size were 1) guilt about eating (EDE-Q Q20, Cohen’s *d*=-0.66) followed by 2) weight importance (EDE-Q Q22, Cohen’s *d*=-0.64), 3) shape importance (EDE-Q Q23, Cohen’s *d*=-0.58), 4) fear of losing control over eating (EDE-Q Q9, Cohen’s *d*=-0.50), and 5) shape/weight preoccupation (EDE-Q Q8, Cohen’s *d*=-0.50) (Figure 1A). Turning to comorbid depressive symptoms, item-level PHQ-9 analysis revealed the greatest overall reductions in appetite/eating concerns (Q5; *d*=−0.51), self-worth (Q6; *d*=−0.30), and anhedonia (Q1; *d*=−0.26) (Figure 1B).

**Figure 1.**
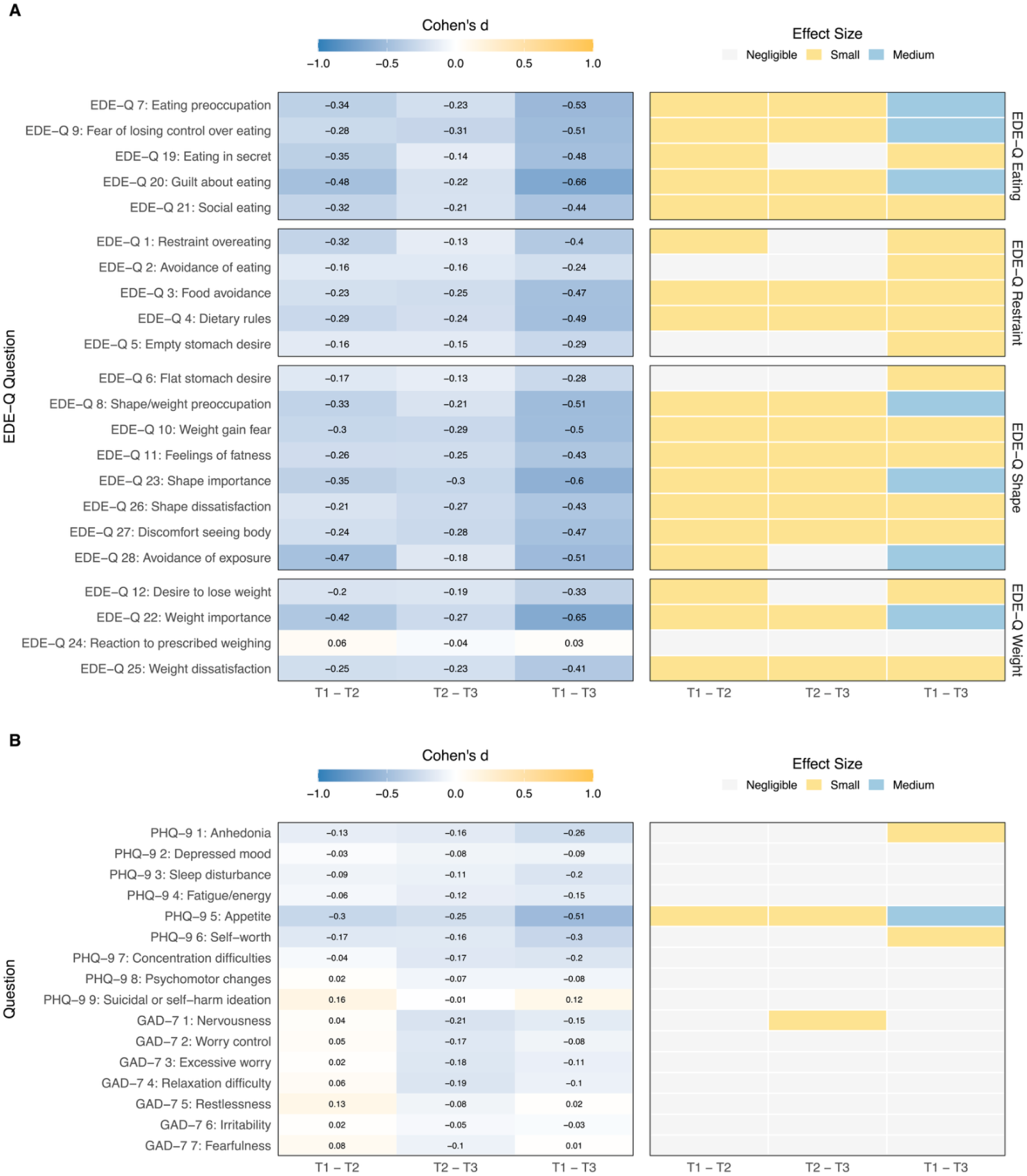
Granular symptom analysis identifies specific targets of the Recovery Record intervention. d=Cohen’s effect size, EDE-Q=Eating Disorder Examination Questionnaire, PHQ-9=Patient Health Questionnaire-9, GAD-7=Generalized Anxiety Disorder 7-item Scale, T=Timepoint **A:** Individual EDE-Q item changes across eating, restraint, shape, and weight concern subscales. Of note, EDE-Q Q8 (shape/weight preoccupation) is part of both the Shape and Weight subscales but was just presented as part of the Shape subscale since it appears first. **B:** Individual PHQ-9 depression and GAD-7 anxiety item changes. Cohen’s d effect sizes are shown for three intervals: early treatment phase (Timepoint 1 [baseline, day 0] to Timepoint 2 [day 15]), late treatment phase (Timepoint 2 to Timepoint 3 [day 30]), and overall treatment (Timepoint 1 to Timepoint 3). Left panels display numeric effect size values with blue-to-yellow gradient indicating magnitude; right panels categorize effects as negligible (|d| < 0.2, white/pale yellow), small (0.2 ≤ |d| < 0.5, yellow), or medium (|d| ≥ 0.5, blue) based on Cohen’s conventions. Negative values indicate symptom reduction (improvement). Eating subscale items showed the largest improvements, with eating preoccupation, fear of losing control over eating, and eating guilt demonstrated medium effect sizes overall. Most improvement occurred during the early treatment phase (Timepoint 1 to Timepoint 2) for eating disorder symptoms.

### Symptom-level changes correlate with overall treatment response

To identify the mechanism of which eating disorder symptoms drive overall treatment response, we examined correlations between EDE-Q Global changes and individual symptom changes from baseline to endpoint, focusing on six symptoms that demonstrated the largest effect sizes in Figure 1 (eating preoccupation [Q7], shape/weight preoccupation [Q8], fear of losing control over eating [Q9], guilt about eating [Q20], weight importance [Q22], shape importance [Q23]) plus one symptom with negligible population-level change (reaction to prescribed weighing [Q24]) as a negative control (Figure 2A and Table S1). Among eating disorder symptoms, shape/weight preoccupation (Q8; *r*=0.66), eating preoccupation (Q7; *r*=0.65), and fear of losing control over eating (Q9; *r*=0.65) showed the strongest correlations with EDE-Q Global score improvement (Figure 2B). In contrast, our negative control symptom, reaction to prescribed weighing (Q24), showed a weaker correlation (*r*=0.38) as hypothesized (Figure 2B). These patterns suggest that improvements in eating preoccupation, shape/weight preoccupation, and fear of losing control over eating are central to overall treatment response.

**Figure 2.**
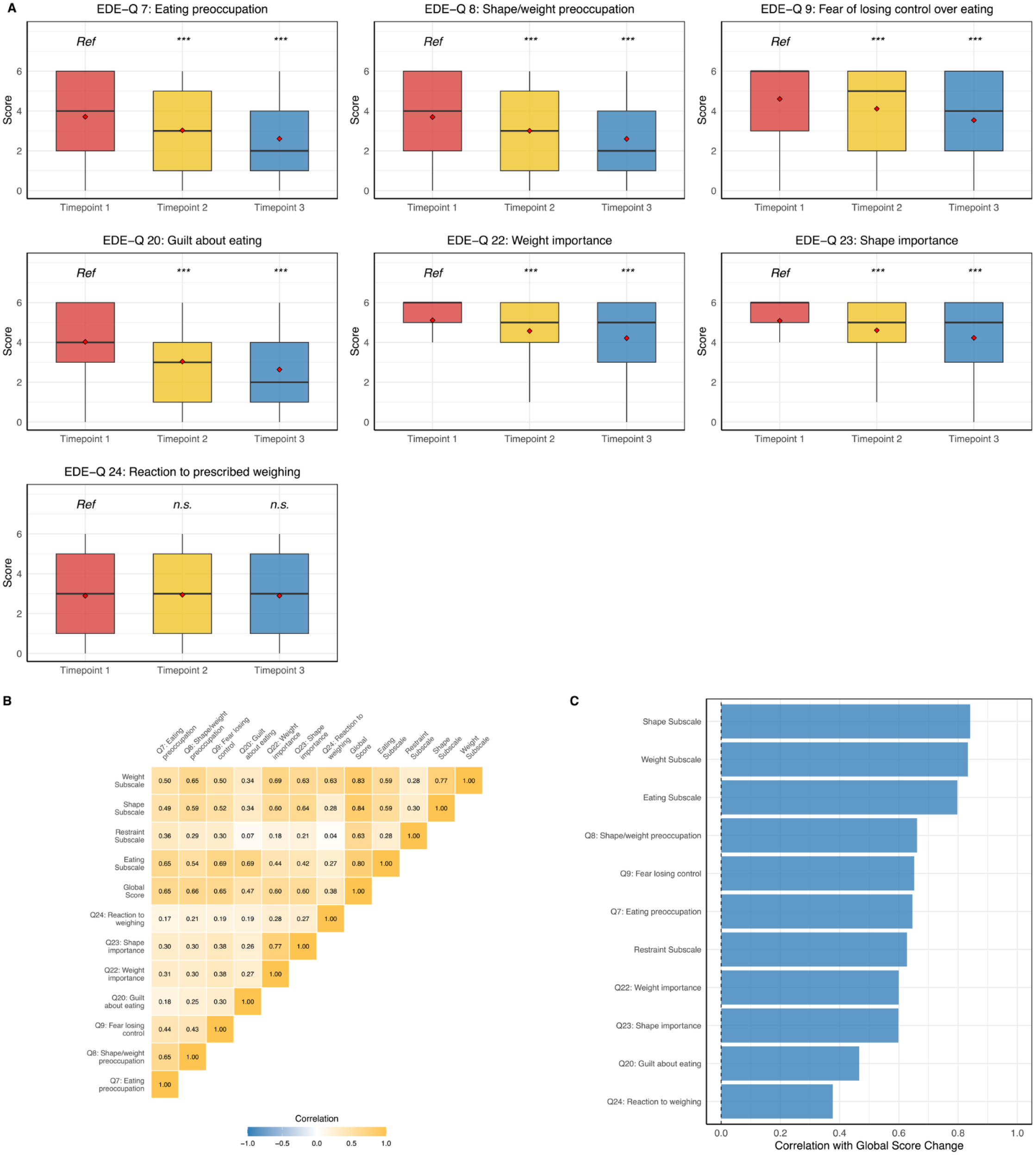
Item-level symptom dynamics identify targets of treatment response. Ref=reference, ns=not significant, *=P<0.05, **=P<0.01, ***=P<0.001, EDE-Q=Eating Disorder Examination Questionnaire **A:** Individual EDE-Q item scores across Timepoint 1 (baseline, day 0), Timepoint 2 (day 15), and Timepoint 3 (day 30). Data are presented as box plots indicating median (center line) and interquartile range (box). **B:** Correlation matrix of the change in individual EDE-Q item scores, subscale scores, and the EDE-Q Global score. **C:** The correlation coefficient between the change in EDE-Q Global score and change in individual EDE-Q questions and EDE-Q subscale scores. The change in eating preoccupation (EDE-Q Q7), shape/weight preoccupation (EDE-Q Q8), and fear of losing control over eating (EDE-Q Q9) were the top three symptoms most correlated with the change in the EDE-Q Global score.

### Baseline symptom levels predict individual treatment response and early symptom improvement mediates treatment response

We next tested whether elevated baseline levels of our three identified symptoms (eating preoccupation [Q7], shape/weight preoccupation [Q8], and fear of losing control over eating [Q9]) could predict individuals most likely to respond to Recovery Record. Individuals with high baseline eating preoccupation (Q7; mean difference=-0.35 points, *P*<0.001), shape/weight preoccupation (Q8; difference=-0.27, *P*<0.001), and fear of losing control over eating (Q9; difference=-0.34, *P*<0.001) demonstrated significantly greater EDE-Q Global score reductions compared to those with low baseline levels of each symptom (Figure 3A). Notably, reaction to prescribed weighing (Q24) showed no sample-level change (Figure 1A) but predicted individual treatment response when elevated at baseline (difference=-0.21, *P*=0.002), indicating heterogeneous response patterns (Figure 3A). All other symptoms showed either weaker or non-significant baseline prediction effects after correction for multiple comparisons.

To test whether these symptoms represent active therapeutic mechanisms rather than merely correlated outcomes, we examined temporal mediation: does early improvement in specific symptoms (Timepoint 1 to Timepoint 2) predict continued EDE-Q Global improvement in the later intervention phase (Timepoint 2 to Timepoint 3)? Linear regression models controlling for baseline EDE-Q Global severity revealed that all examined symptoms showed significant temporal mediation effects (all *P*<0.001, Bonferroni-corrected) (Figure 3B). Weight importance (Q22) demonstrated the strongest mediation coefficient (β=-0.11, *P*<0.001), followed by shape/weight preoccupation (Q8; β=-0.10), shape importance (Q23; β=-0.09), eating preoccupation (Q7; β=-0.09), fear of losing control over eating (Q9; β=-0.08), guilt about eating (Q20; β=-0.08), and reaction to prescribed weighing (Q24; β=-0.06) (Figure 3B). These findings indicate that early symptom changes predict sustained improvement trajectories, supporting these parameters as relevant mechanisms in the treatment of binge-type eating disorders using Recovery Record.

**Figure 3.**
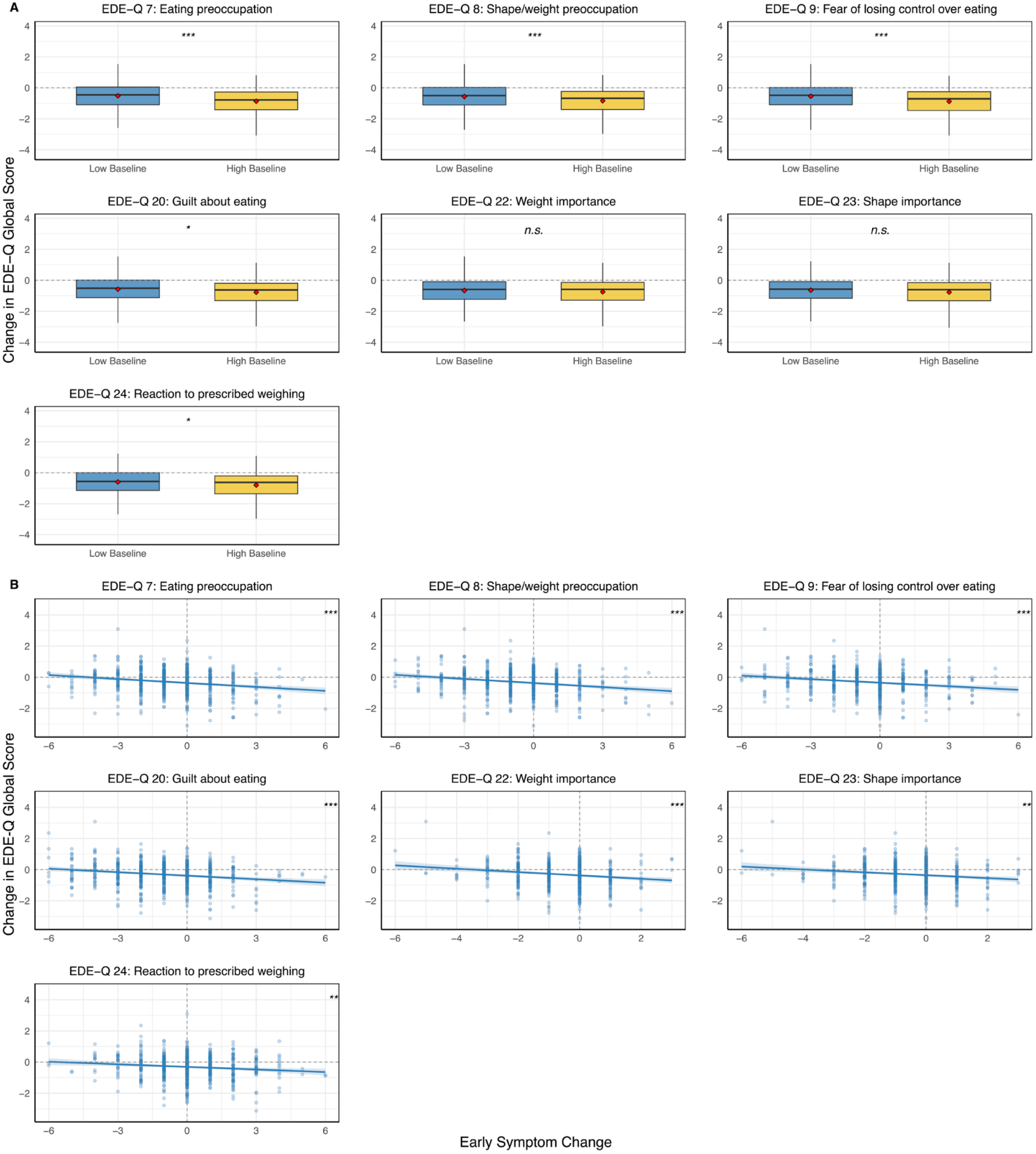
Baseline symptom severity and early symptom change identify symptom-specific drivers of treatment response. Ref=reference, ns=not significant, *=P<0.05, **=P<0.01, ***=P<0.001, EDE-Q=Eating Disorder Examination Questionnaire **A**: Distribution of changes in EDE-Q Global scores among individuals with low versus high baseline EDE-Q item-level scores, defined by a median split. **B:** Correlation between the change in EDE-Q Global scores and the change in EDE-Q item level scores (Timepoint 1 to Timepoint 2). Individuals with higher baseline severity of eating preoccupation (EDE-Q Q7), shape/weight preoccupation (EDE-Q Q8), and fear of loss of control over eating (EDE-Q Q9) demonstrated greater treatment-related improvements in EDE-Q Global scores, with symptom improvement emerging between Timepoint 1 and Timepoint 2.

### Cluster analysis reveals distinct response profiles defined by baseline symptom presentation

To identify naturally occurring response subgroups, we performed hierarchical clustering on individual symptom change trajectories for EDE-Q Global scores and examined the corresponding change in eating preoccupation (Q7), shape/weight preoccupation (Q8), and fear of losing control over eating (Q9). We evaluated clustering solutions using three complementary approaches: the elbow method, silhouette analysis, and gap statistic. While silhouette scores indicated weak-to-moderate cluster separation (average width=0.26), the elbow method identified k=3 as the optimal solution balancing parsimony with clinical interpretability (Figure S3). The gap statistic showed maximum values at k=1, suggesting continuous rather than discrete response patterns, but k=3 provided meaningful clinical distinctions (Figure S3).

Hierarchical clustering revealed three distinct response profiles (Figure 4A and 4B). Non-responders (n=129, 18.4%) demonstrated minimal change (EDE-Q Global: M=-0.06, SD=0.75) with slight worsening on eating preoccupation (Q7: M=1.11). Moderate responders (n=326, 46.4%) showed modest improvements (EDE-Q Global: M=-0.36, SD=0.53; eating preoccupation (Q7): M=-0.69; shape/weight preoccupation (Q8): M=-0.73; fear of losing control over eating (Q9): M=0.24). Strong responders (n=247, 35.2%) achieved substantial improvements across all measures (EDE-Q Global: M=-1.50, SD=0.70; eating preoccupation (Q7): M=-2.55; shape/weight preoccupation (Q8): M=-2.41; fear of losing control over eating (Q9): M=-2.93) (Figure 4B).

Critically, response clusters differed significantly in baseline symptom presentation. Non-responders showed significantly lower baseline eating preoccupation (Q7: mean=2.36) compared to moderate responders (mean=3.75) and strong responders (mean=4.17, *P*<0.001) (Figure 4C). Similar patterns emerged for shape/weight preoccupation (Q8: *P*<0.001), fear of losing control over eating (Q9: *P*<0.001), and EDE-Q Global severity (*P*<0.001) (Figure 4C). In contrast, reaction to prescribed weighing (Q24) did not differ significantly across clusters (*P*=0.53) (Figure 4C). These findings suggest that individuals presenting with elevated levels of eating preoccupation, shape/weight preoccupation, and fear of losing control over eating at baseline may be more likely to respond to Recovery Record, and these symptoms may serve as precision psychological markers.

**Figure 4.**
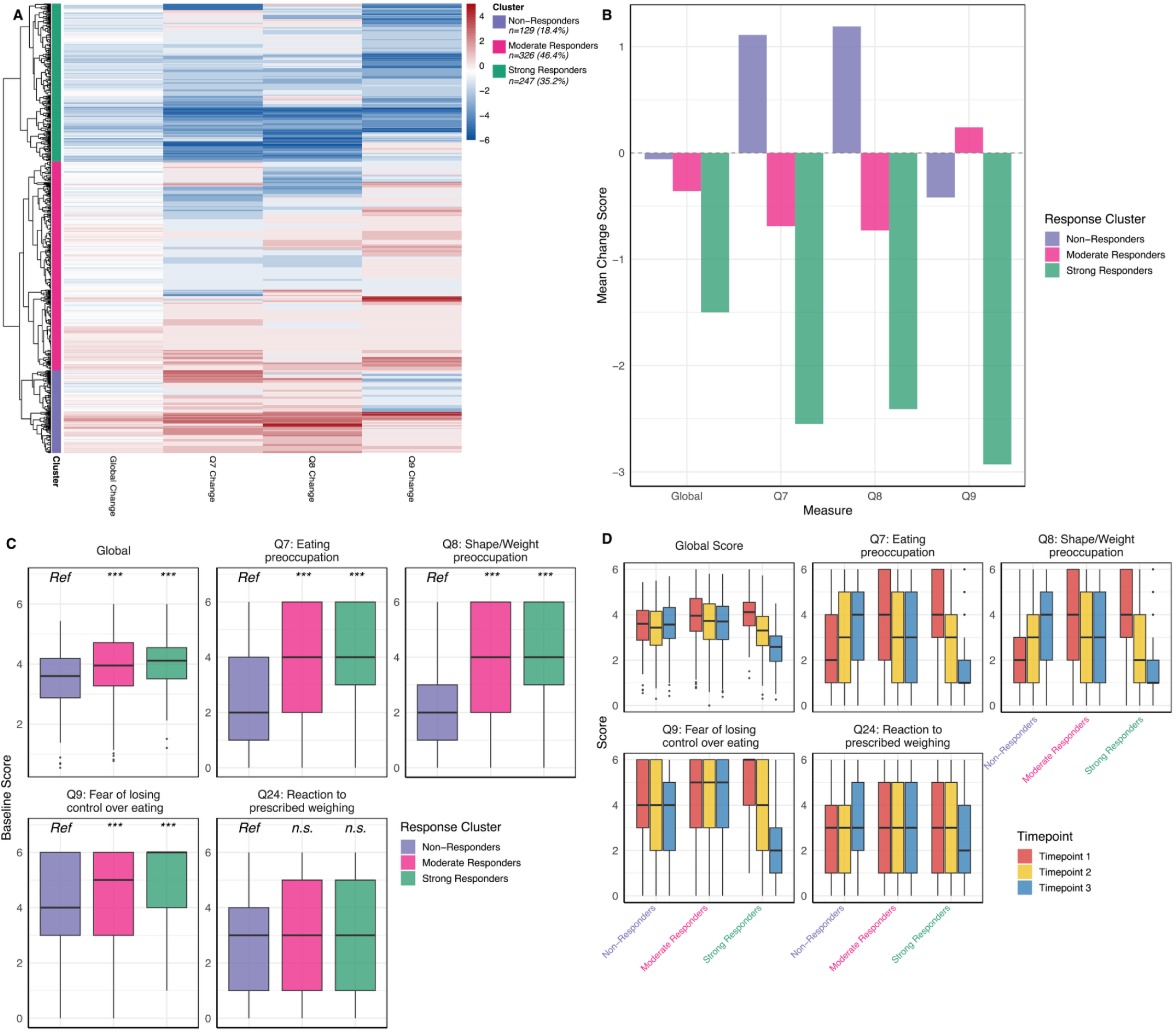
Recovery Record users cluster into three treatment response groups associated with distinct symptom profiles. Ref=reference, ns=not significant, *=P<0.05, **=P<0.01, ***=P<0.001, EDE-Q=Eating Disorder Examination Questionnaire **A:** Treatment response clusters of Recovery Record users. Clustering was performed using changes in EDE-Q Global scores between Timepoint 1 and Timepoint 3, with corresponding changes in eating preoccupation (EDE-Q Q7), shape/weight preoccupation (EDE-Q Q8), and fear of loss of control over eating (EDE-Q Q9) displayed to highlight associations with EDE-Q Global treatment response. **B:** Mean change in EDE-Q Global, eating preoccupation (EDE-Q Q7), shape/weight preoccupation (EDE-Q Q8), and fear of loss of control over eating (EDE-Q Q9) among non-responders, moderate responders, and strong responders of the Recovery Record intervention. **C:** Baseline levels of EDE-Q Global, eating preoccupation (EDE-Q Q7), shape/weight preoccupation (EDE-Q Q8), and fear of loss of control over eating (EDE-Q Q9), and reaction to prescribed weighing (EDE-Q Q24, control) among non-responders, moderate responders, and strong responders of the Recovery Record intervention. **D:** Scores of EDE-Q Global, eating preoccupation (EDE-Q Q7), shape/weight preoccupation (EDE-Q Q8), and fear of loss of control over eating (EDE-Q Q9), and reaction to prescribed weighing (EDE-Q Q24, control) across Timepoint 1 (day 0), Timepoint 2 (day 15), and Timepoint 3 (day 30) among non-responders, moderate responders, and strong responders of the Recovery Record intervention. Eating preoccupation (EDE-Q Q7), shape/weight preoccupation (EDE-Q Q8), and fear of loss of control over eating (EDE-Q Q9) function as precision psychological markers of the Recovery Record intervention.

### Response clusters show similar demographic profiles with notable diagnostic differences

We characterized demographic and clinical characteristics across the three response clusters to determine whether response patterns reflected distinct patient subgroups. Surprisingly, response clusters demonstrated remarkably similar demographic profiles. Sex distribution did not differ across clusters (86.8% female in non-responders, 88.0% in moderate responders, and 89.9% in strong responders; *P*=0.64), nor did age (means: 29.5, 30.1, and 30.3 years respectively; *P*=0.62) or racial/ethnic composition (*P*=0.89), or current BMI (Figures S4A-C).

Among clinical features, compensatory behaviors differed across clusters (global *P*=0.037), driven primarily by differences in diet pill use, with non-responders reporting the highest rates (55.8% in non-responders, 39.8% in moderate responders, and 47.4% in strong responders; *P*<0.01; Figure 5A). Lifetime eating disorder diagnosis showed differences across clusters (global *P*=0.042), though no pairwise comparison survived Bonferroni adjustment (Figure 5B). Family history of eating disorders increased across response clusters, with 10.1% of non-responders, 17.2% of moderate responders, and 20.2% of strong responders reporting a biological relative with an eating disorder (global *P*=0.046), with strong responders significantly more likely than non-responders to report a family history (*P*=0.037; Figure 5C). We did not observe any differences in response clusters by current weight, treatment type, highest BMI, lowest adult BMI, age at menarche (among females), age at first binge, or age at last binge (i.e., age at enrollment) (Figures S4D-S4I).

**Figure 5.**
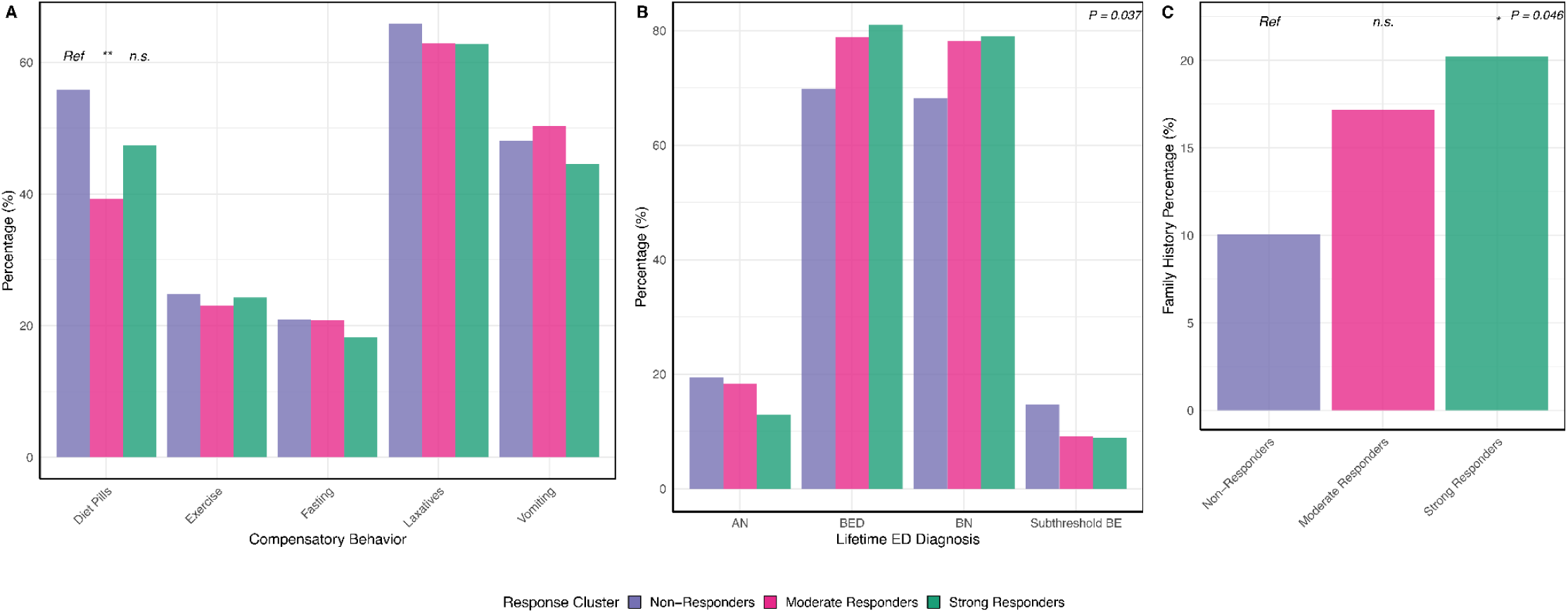
Demographic and clinical characteristics in Recovery Record response clusters. Ref=reference, ns=not significant, *=P<0.05, **=P<0.01, ***=P<0.001, ED=eating disorder, AN=anorexia nervosa, BED=binge-eating disorder, BN=bulimia nervosa, BE=binge eating. The proportion of individuals among Recovery Record non-responders, moderate responders, and strong responders by compensatory behavior (A), lifetime eating disorder diagnosis (B), and family history (C). Global P-values are reported for each panel. Only features exhibiting at least one significant difference among non-responders, moderate responders, and strong responders are labeled with significance annotations; all others were not significantly different relative to non-responders.

### Convergent evidence identifies three precision psychological markers

Across four complementary analytical approaches (correlation analysis, baseline prediction, temporal mediation, and cluster differentiation), eating preoccupation (Q7), shape/weight preoccupation (Q8), and fear of losing control over eating (Q9) consistently demonstrated the strongest and most convergent evidence as precision psychological markers for Recovery Record treatment response (Figure 6). These three symptoms showed the strongest correlations with overall EDE-Q Global improvement, predicted greater response when elevated at baseline, mediated sustained therapeutic gains through early symptom change, and differentiated naturally occurring response clusters. In contrast, guilt about eating (Q20) and weight importance (Q22) met correlation and mediation criteria but did not differentiate response clusters, and shape importance (Q23) met only the correlation criterion. Reaction to prescribed weighing (Q24), included as a negative control, showed weaker and inconsistent evidence across criteria and did not differentiate response clusters, supporting the overall specificity of the framework.

**Figure 6.**
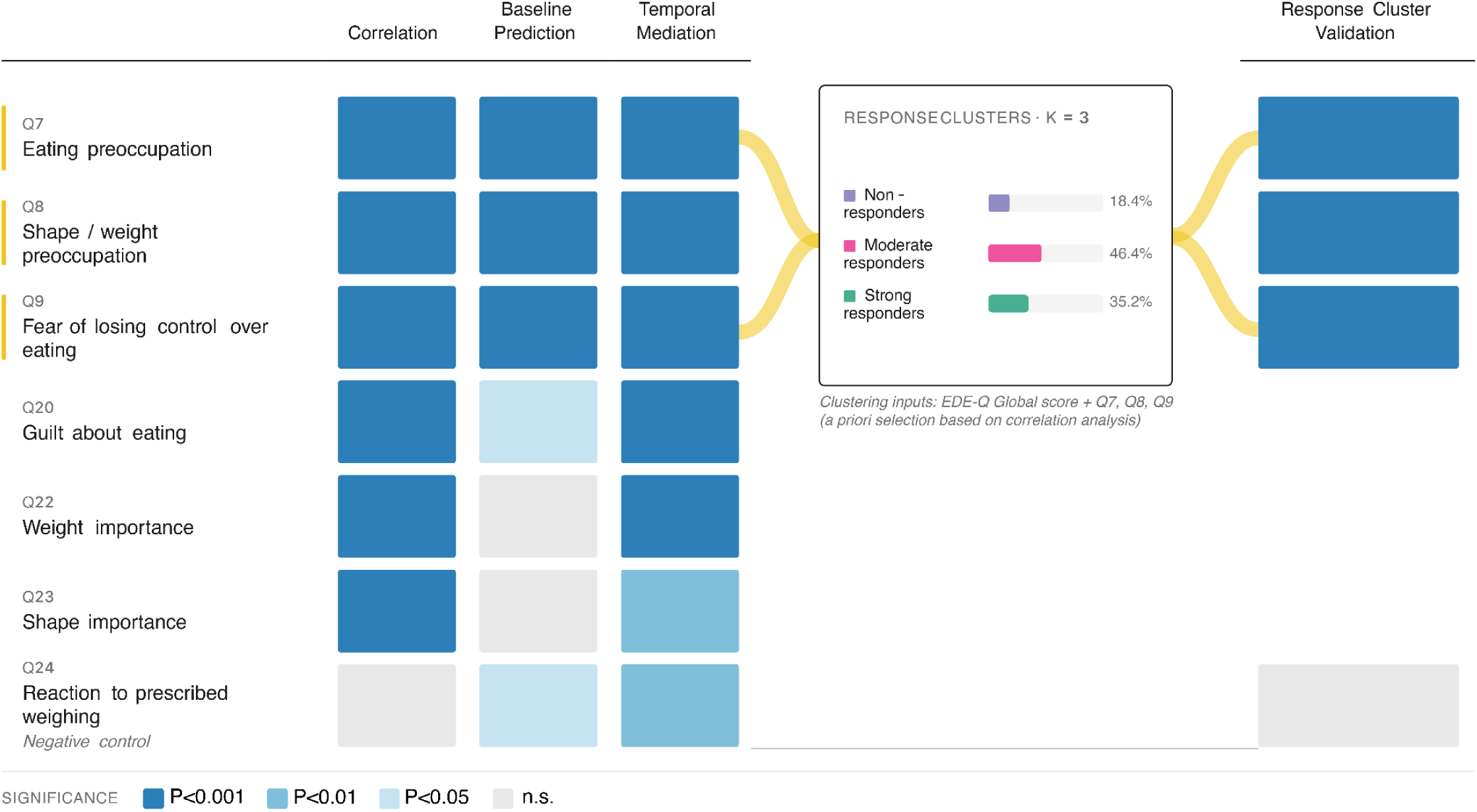
Four-criterion convergent framework identifies precision psychological markers for Recovery Record treatment response. EDE-Q=Eating Disorder Examination Questionnaire, ns=not significant Seven candidate EDE-Q symptoms (rows) are evaluated across four analytical criteria (columns): correlation analysis, baseline prediction, temporal mediation, and response cluster validation. Cell shading reflects statistical significance: dark blue (P<0.001), medium blue (P<0.01), light blue (P<0.05), and grey (n.s.). Q7, Q8, and Q9 (yellow left border) met criteria across all four approaches and were designated precision psychological markers. Yellow ribbons indicate that EDE-Q Global score change and Q7, Q8, and Q9 change scores were selected a priori as clustering inputs based on correlation analysis findings, with response cluster validation confirming their differentiation of non-responders (18.4%), moderate responders (46.4%), and strong responders (35.2%). Q24 (reaction to prescribed weighing) is included as a negative control.

## Discussion

This study provides the first comprehensive mechanistic analysis of Recovery Record for eating disorders, demonstrating significant symptom improvements over 30 days and revealing substantial heterogeneity in treatment response. Through converging evidence from correlation analysis, baseline prediction, temporal mediation, and cluster analysis, we identified eating preoccupation (Q7), shape/weight preoccupation (Q8), and fear of losing control over eating (Q9) as precision psychological markers for Recovery Record response. Individuals with elevated baseline levels of these symptoms were more likely to achieve substantial benefit, with 35% achieving strong response compared to 18% showing minimal response. These findings are consistent with Tregarthen et al., who reported 61.6% of users showed clinically meaningful change16; our mechanistic analyses extend this work by revealing substantial heterogeneity within the responding population, with strong responders achieving over four-fold greater improvement than moderate responders.

The identification of these symptoms as precision psychological markers represents a meaningful step toward targeted digital mental health care. Baseline assessment of eating preoccupation, shape/weight preoccupation, and fear of losing control over eating can prospectively identify individuals most likely to respond to Recovery Record, paralleling precision medicine approaches where biomarker-driven patient selection has transformed treatment^12^. Strong responders presented with significantly higher baseline levels of all three symptoms compared to non-responders, suggesting that individuals lacking these elevated symptom profiles at baseline may be suboptimal candidates for standard Recovery Record content. The temporal mediation findings further support these symptoms as active therapeutic mechanisms rather than merely correlated outcomes: early improvements in these symptoms (Timepoint 1 to Timepoint 2) predicted sustained EDE-Q Global improvement in the later phase (Timepoint 2 to Timepoint 3), consistent with a mechanistic role in the intervention’s therapeutic process. The temporal dynamics observed (rapid initial improvement followed by more modest continued gains) are consistent with CBT research, where accelerated early change likely reflects both therapeutic expectancy effects and the immediate impact of self-monitoring^40,41^. Notably, the magnitude of EDE-Q Global improvement observed across all Recovery Record users (∼0.9 points) was comparable to that achieved following 16 weeks of structured group CBT for BN and BED, with strong responders exceeding this benchmark (∼1.7 points) and approaching the established full remission threshold of ≤2.35, a clinically meaningful signal achieved in 30 days without therapist involvement^42,43^.

The precision marker symptoms reflect the centrality of cognitive preoccupation and control concerns in eating disorder maintenance. Eating preoccupation consumes mental resources and drives compensatory behaviors; shape/weight preoccupation fuels appearance-based self-evaluation and body monitoring; and fear of losing control reinforces rigid dietary rules that paradoxically increase the risk of engaging in a binge. Individuals whose disorders are dominated by these cognitive processes appear to have the greatest capacity to benefit from CBT-based strategies delivered digitally.

In contrast, guilt about eating (Q20) and weight importance (Q22) demonstrated large population-level improvements and strong temporal mediation effects but did not differentiate response clusters, indicating they operate as universal therapeutic mechanisms that benefit most users regardless of baseline presentation. This distinction has important implications for intervention design: universal mechanisms represent core therapeutic targets for all users, while precision markers identify opportunities for personalized content delivery; for instance, routing individuals with low precision marker scores but elevated guilt about eating toward alternative intervention modules within Recovery Record. Together, these findings suggest Recovery Record’s effectiveness stems from two complementary processes: precision targeting of modifiable cognitive-behavioral mechanisms in susceptible individuals, and broadly accessible therapeutic pathways that support recovery across the wider user population.

Response clusters demonstrated remarkably similar demographic profiles, strengthening the conclusion that response variability was driven by specific symptom presentations rather than patient characteristics. Among clinical features, diet pill use showed a U-shaped pattern across clusters (highest in non-responders, lower in moderate responders, rising again in strong responders), potentially reflecting heterogeneous underlying motivations or illness severity profiles. Response cluster distributions in BED diagnosis showed significant overall group differences, though pairwise comparisons did not survive Bonferroni correction; however, strong responders were significantly more likely than non-responders to report a family history of eating disorders after Bonferroni correction, suggesting meaningful clinical variation across response groups that warrants further investigation in larger samples.

While greater app engagement was associated with improvements in EDE-Q Global and Shape Concern scores, most outcomes showed no significant engagement effect, and confidence intervals were wide across all measures^44^. Prior work from this cohort has characterized engagement with specific app components and modeled how engagement changes over time; yet how engagement patterns intersect with symptom-based response profiles remains unexamined^18,45^. A natural extension of the current precision marker framework is to incorporate engagement dimensions alongside clinical markers.

Our findings support implementing a two-stage precision framework for Recovery Record and for digital mental health interventions more broadly. In Stage 1 (Baseline Assessment), prospective users complete a brief assessment of precision psychological marker symptoms prior to beginning the intervention. Individuals presenting with elevated baseline levels represent optimal candidates; those below empirically-derived thresholds should be directed toward adapted intervention pathways within Recovery Record. Given persistent barriers to traditional care and limited evidence guiding next steps for CBT non-responders in eating disorders, app-based adaptations represent a scalable and clinically meaningful path forward^46,47^. In Stage 2 (Early Response Monitoring), users are reassessed at a midpoint timepoint on the same precision marker symptoms. Early improvement predicts sustained response and validates continued engagement with the current approach; absence of early improvement may signal need for intensification or transition to alternative modules^47^. Because precision markers are discrete, quantifiable symptom scores, triage and monitoring workflows could be algorithmically embedded directly into digital platforms, reducing clinician burden while maintaining clinical oversight.

The timing of this framework is particularly opportune. In December 2025, the FDA launched TEMPO (Testing and Evidence for Digital Health Optimization), a first-of-its-kind pilot enabling Medicare beneficiaries to access FDA-authorized digital therapeutic devices through streamlined pathways prioritizing real-world evidence generation^48^. As digital mental health interventions integrate into frameworks like TEMPO and value-based care models, precision psychological marker algorithms will be essential for demonstrating appropriate patient selection and maximizing cost-effectiveness^49^.

Several limitations warrant consideration. First, the sample was predominantly female, White non-Hispanic, and young adults, limiting generalizability; although response clusters did not differ significantly by demographic characteristics in the current sample, precision psychological markers may operate differently when investigated in larger groups of underrepresented populations^41^. Second, the observational design precludes causal conclusions about intervention effects, as improvements could reflect natural recovery, regression to the mean, or other temporal factors, though temporal mediation analyses support intervention-driven change. Third, BEGIN was designed to identify predictors of binge eating rather than as a treatment trial, and intervention effects should be interpreted accordingly; duration of illness data were not collected, precluding examination of this potentially important moderator. Fourth, elevated baseline symptom levels predicting greater treatment response may partially reflect floor and ceiling effects, where individuals with lower baseline severity have less room to improve; although including baseline EDE-Q Global severity as a covariate partially accounts for this, it cannot be fully ruled out. Lower baseline scores may also reflect prior treatment, differences in illness trajectory, or prior Recovery Record exposure rendering intervention content less novel, which are possibilities that cannot be disentangled as treatment history, illness duration, and previous app use were not assessed. Fifth, we operationalized engagement as total app interaction counts, an aggregate metric that may obscure meaningful differences in engagement quality and feature-specific use.

Prospective validation of the eating preoccupation (EDE-Q Q7), shape/weight preoccupation (EDE-Q Q8), and fear of losing control over eating (EDE-Q Q9) precision marker framework in independent samples is essential before clinical implementation, ideally through a randomized trial stratifying participants by baseline marker levels and comparing Recovery Record to alternative pathways. Investigating why individuals with elevated eating preoccupation, shape/weight preoccupation, and fear of losing control over eating respond more robustly (whether due to Recovery Record’s specific feature set, greater motivation for change, or differential symptom plasticity) would inform targeted intervention optimization. Expanding investigation to underrepresented populations and integrating cost-effectiveness analyses comparing precision marker-guided versus universal access will be critical for sustainable implementation at scale.

This study demonstrates that Recovery Record produces significant improvements in eating disorder symptoms while revealing substantial heterogeneity in treatment response. Eating preoccupation, shape/weight preoccupation, and fear of losing control over eating function as precision psychological markers that prospectively identify individuals most likely to benefit from Recovery Record. Early improvements in these symptoms further mediate sustained therapeutic gains, supporting their role as active mechanisms of intervention response. As digital interventions become increasingly integrated into mental healthcare systems through initiatives like FDA TEMPO and value-based payment models, precision psychological marker frameworks will be essential for optimizing resource allocation, maximizing effectiveness, and ensuring digital tools reach the populations most likely to benefit.

## Acknowledgements

We extend our sincere gratitude to all BEGIN participants whose contributions made this research possible. This work was supported by the National Science Foundation (DGE-1650116; REF, principal investigator); Foundation of Hope, Raleigh, North Carolina (CMB, principal investigator); National Eating Disorders Association (CMB and JT, principal investigators); Brain and Behavior Research Foundation (NARSAD Distinguished Investigator Grant; CMB, principal investigator); and the National Institute of Mental Health (R01MH119084, CMB/Butner, MPIs; U01MH109528, Sullivan principal investigator, CMB co-investigator). The views and conclusions expressed herein are those of the authors alone and do not represent the positions of the National Science Foundation. Apple Inc. provided the Apple Watch devices for the research. Apple was not involved in the design of the research, nor was it involved in the collection, analysis, or interpretation of the research data, or the content of this or any related publication.

## Funding

CMB is supported by NIMH (R01MH136149; R01MH134039; R56MH129437; R01MH124871; R01MH124871). EH and MAH are supported by NHGRI (RM1HG010860).

## Data availability

Data generated or analyzed in this study are not publicly available given proprietary restrictions designated by Recovery Record, but may be requested from the corresponding author.

## Author contributions

EH conceived and designed the analytic study, while LMT and CMB designed the BEGIN study. EH and LM analyzed the data. EH, LM, REF, KKR, ZB, MAH, LMT, CMB, and RP interpreted the results. EH wrote the draft manuscript. MAH, RP, LMT, and CMB provided supervision. MAH and CMB acquired funding. All authors reviewed and edited the final manuscript for publication.

## Notes

### Competing Interest Statement

The authors have declared no competing interest.

### Author Declarations

The Institutional Review Board of University of North Carolina gave ethical approval for data collection in the BEGIN study (Protocol #17-0242).

