## Supplementary material for "Precision psychological markers enable targeted treatment in digital eating disorder interventions"

### Supplementary results

**Table S1. EDE-Q questions and symptoms**

| Question number | Question | Subscale | Symptom |
|---|---|---|---|
| 1 | Have you been deliberately trying to limit the amount of food you eat to influence your shape or weight (whether or not you have succeeded)? | Restraint | Restraint overeating |
| 2 | Have you gone for long periods of time (8 waking hours or more) without eating anything at all in order to influence your shape or weight? | Restraint | Avoidance of eating |
| 3 | Have you tried to exclude from your diet any foods that you like in order to influence your shape or weight (whether or not you have succeeded)? | Restraint | Food avoidance |
| 4 | Have you tried to follow definite rules regarding your eating (for example, a calorie limit) in order to influence your shape or weight (whether or not you have succeeded)? | Restraint | Dietary rules |
| 5 | Have you had a definite desire to have an empty stomach with the aim of influencing your shape or weight? | Restraint | Empty stomach desire |
| 6 | Have you had a definite desire to have a totally flat stomach? | Shape | Flat stomach desire |
| 7 | Has thinking about food, eating or calories made it very difficult to concentrate on things you are interested in (for example, working, following a conversation, or reading)? | Eating | Eating preoccupation |
| 8 | Has thinking about shape or weight made it very difficult to concentrate on things you are interested in (for example, working, following a conversation, or reading)? | Shape and Weight | Shape/weight preoccupation |
| 9 | Have you had a definite fear of losing control over eating? | Eating | Fear of losing control over eating |
| 10 | Have you had a definite fear that you might gain weight? | Shape | Weight gain fear |
| 11 | Have you felt fat? | Shape | Feelings of fatness |
| 12 | Have you had a strong desire to lose weight? | Weight | Desire to lose weight |

| | | | |
|---|---|---|---|
| 13 | Over the past 28 days, how many times have you eaten what other people would regards as an unusually large amount of food (given the circumstances)? | Behavioral frequency | Overeating frequency (times) |
| 14 | … On how many of these times did you have a sense of having lost control over your eating (at the time you were eating)? | Behavioral frequency | Overeating loss of control |
| 15 | Over the past 28 days, on how many DAYS have such episodes of overeating occurred (i.e. you have eaten an unusually large amount of food and have had a sense of loss of control at the time)? | Behavioral frequency | Overeating frequency (days) |
| 16 | Over the past 28 days, how many times have you made yourself sick (vomit) as a means of controlling your shape or weight? | Behavioral frequency | Self-induced vomiting frequency |
| 17 | Over the past 28 days, how many times have you taken laxatives as a means of controlling your shape or weight? | Behavioral frequency | Laxative frequency |
| 18 | Over the past 28 days, how many times have you exercised in a “driven” or “compulsive” way as a means of controlling your weight, shape or amount of fat, or to burn off calories? | Behavioral frequency | Compulsive exercise frequency |
| 19 | Over the past 28 days, on how many days have you eaten in secret (ie, furtively)? … Do not count episodes of binge eating. | Eating | Eating in secret |
| 20 | On what proportion of the times that you have eaten have you felt guilty (felt that you’ve done wrong) because of its effect on your shape or weight? … Do not count episodes of binge eating. | Eating | Guilt about eating |
| 21 | Over the past 28 days, how concerned have you been about other people seeing you eat? … Do not count episodes of binge eating. | Eating | Social eating |
| 22 | Has your weight influenced how you think about (judge) yourself as a person? | Weight | Weight importance |

| | | | |
|---|---|---|---|
| 23 | Has your shape influenced how you think about (judge) yourself as a person? | Shape | Shape importance |
| 24 | How much would it have upset you if you had been asked to weigh yourself once a week (no more, or less, often) for the next four weeks? | Weight | Reaction to prescribed weighing |
| 25 | How dissatisfied have you been with your weight? | Weight | Weight dissatisfaction |
| 26 | How dissatisfied have you been with your shape? | Shape | Shape dissatisfaction |
| 27 | How uncomfortable have you felt seeing your body (for example, seeing your shape in the mirror, in a shop window reflection, while undressing or taking a bath or shower)? | Shape | Discomfort seeing body |
| 28 | How uncomfortable have you felt about others seeing your shape or figure (for example, in communal changing rooms, when swimming, or wearing tight clothes)? | Shape | Avoidance of exposure |

**Table S2. PHQ-9, GAD-7, and symptom categories**

| Assessment | Question number | Question | Symptom |
|---|---|---|---|
| PHQ-9 | 1 | Little interest or pleasure in doing things | Anhedonia |
| PHQ-9 | 2 | Feeling down, depressed, or hopeless | Depressed mood |
| PHQ-9 | 3 | Trouble falling or staying asleep, or sleeping too much | Sleep disturbance |
| PHQ-9 | 4 | Feeling tired or having little energy | Fatigue/energy |
| PHQ-9 | 5 | Poor appetite or overeating | Appetite |
| PHQ-9 | 6 | Feeling bad about yourself - or feeling like you are a failure or have let yourself or your family down | Self-worth |
| PHQ-9 | 7 | Trouble concentrating on things, such as reading the newspaper or watching television | Concentration difficulties |
| PHQ-9 | 8 | Moving or speaking so slowly that other people could have noticed? Or the opposite - being so fidgety or restless that you have been moving around a lot more than usual | Psychomotor changes |
| PHQ-9 | 9 | Thoughts that you would be better off dead or hurting yourself in some way | Suicidal or self-harm ideation |
| GAD-7 | 1 | Feeling nervous, anxious, or on edge | Nervousness |
| GAD-7 | 2 | Not being able to stop or control worrying | Worry control |
| GAD-7 | 3 | Worrying too much about different things | Excessive worry |
| GAD-7 | 4 | Trouble relaxing | Relaxation difficulty |
| GAD-7 | 5 | Being so restless that it is hard to sit still | Restlessness |
| GAD-7 | 6 | Becoming easily annoyed or irritable | Irritability |
| GAD-7 | 7 | Feeling afraid, as if something awful might happen | Fearfulness |

**Table S3. Assessment completion rates across study visits**

| Visit | Number of people (%) |
|---|---|
| 1 (day 0) | 1,124 (96.4%) |
| 2 (day 15) | 859 (73.7%) |
| 3 (day 30) | 734 (63.0%) |

**Figure S1. Eating disorder symptoms show significant improvement following 30-Day Recovery Record intervention**

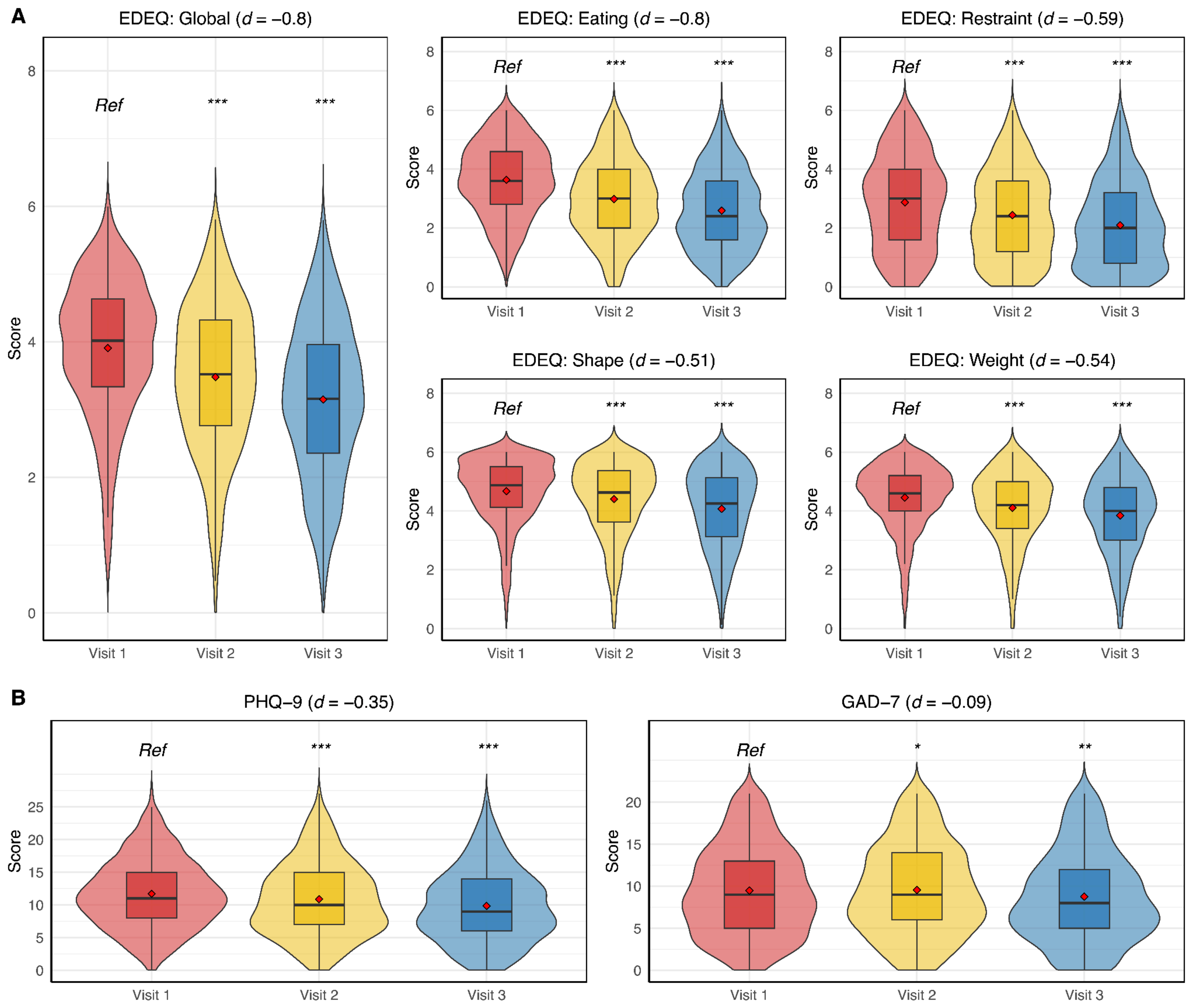


*Ref*=reference, *ns*=not significant, *=*P*<0.05, **= *P*<0.01, ***= *P*<0.001, *d*=Cohen's effect size, EDE-Q=eating disorder examination questionnaire, PHQ-9=patient health questionnaire 9, GAD-7=generalized anxiety disorder 7-item scale

*EDE-Q Global and subscale scores (A) and PHQ-9 depression and GAD-7 anxiety scores (B) at baseline (Timepoint 1, day 0), mid-treatment (Timepoint 2, day 15), and end-of-treatment (Timepoint 3, day 30). Violin plots show score distributions with embedded box plots indicating median (center*

*line) and interquartile range (box). Cohen's d effect sizes (relative to baseline) are shown in panel titles. Linear mixed-effects models compared each timepoint to baseline (Ref). All measures demonstrated significant improvement throughout treatment.*

**Figure S2. Symptom changes as a function of app engagement**

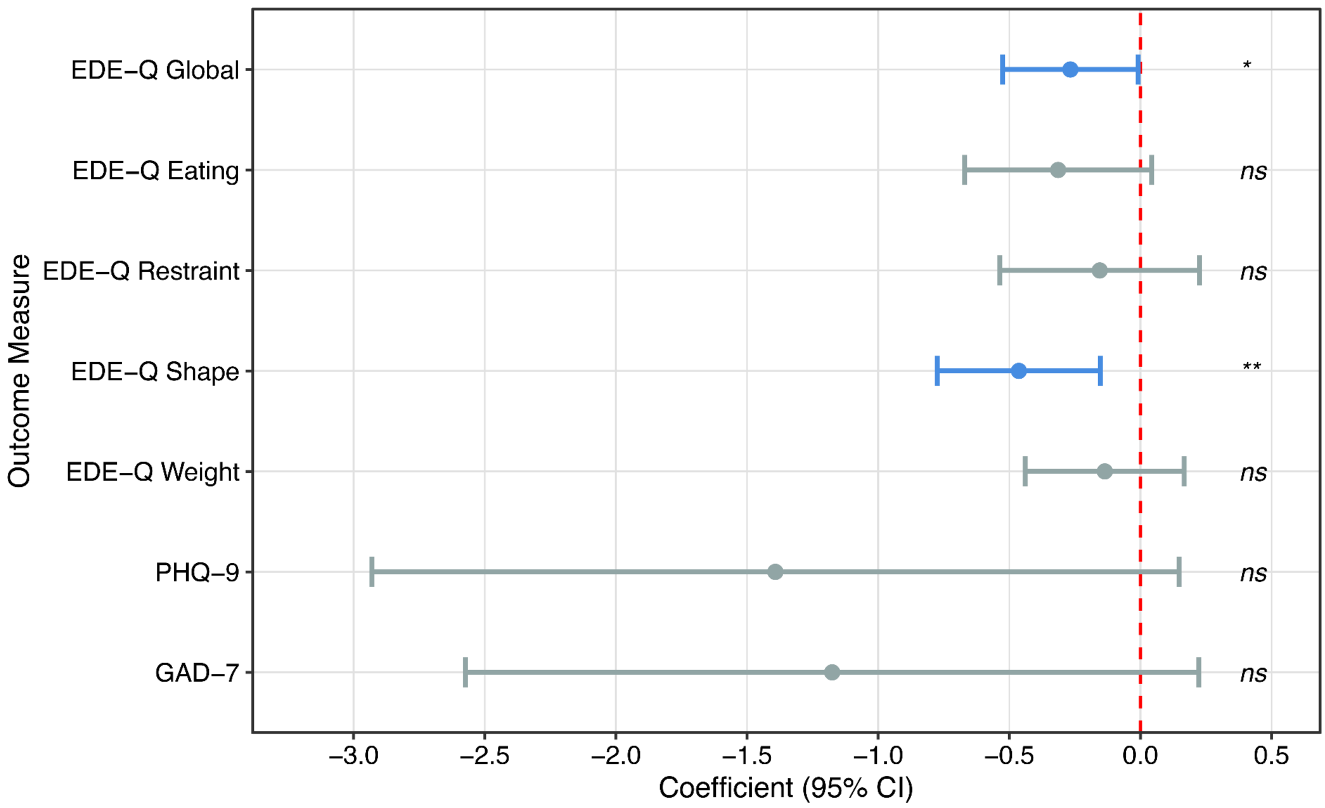


*Ref*=reference, *ns*=not significant, *=*P*<0.05, **=*P*<0.01, ***=*P*<0.001, EDE-Q=eating disorder examination questionnaire, PHQ-9=patient health questionnaire 9, GAD-7=generalized anxiety disorder 7-item scale

*Forest plot of standardized regression coefficients (95% CI) examining continuous engagement count as a predictor of symptom change from baseline (Timepoint 1) to end-of-treatment (Timepoint 3). Linear regression models included age, sex, and race/ethnicity as covariates.*

**Figure S3. Determination of the optimal number of treatment response clusters**

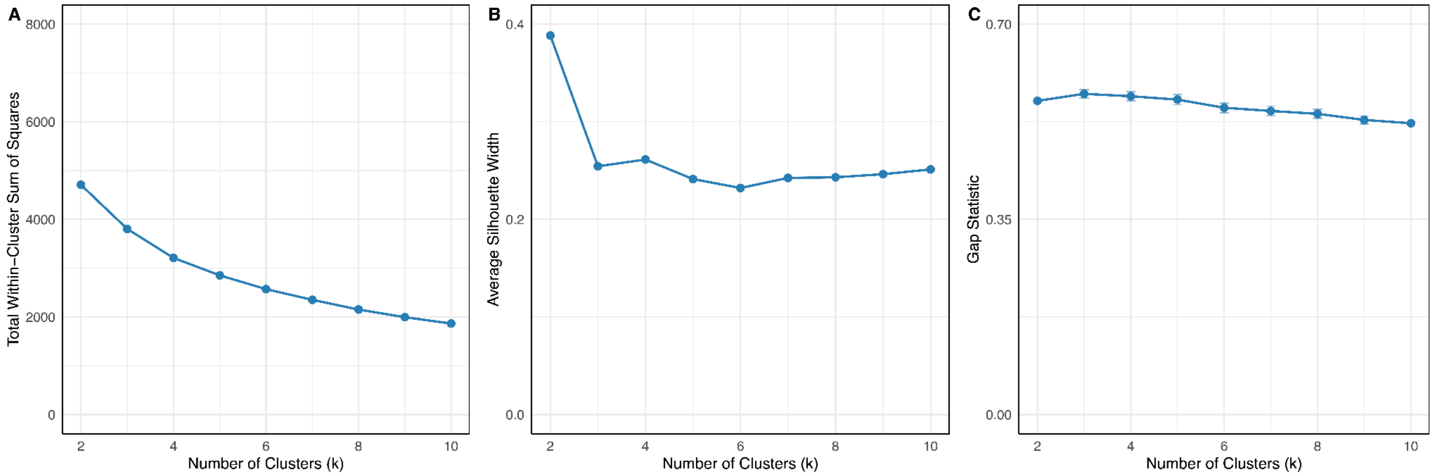


*Quantifying the number of clusters in Recovery Record users using the elbow method (A), silhouette score (B), and (C). Collectively, these metrics support a three-cluster solution among Recovery Record users.*

**Figure S4. Additional demographic and clinical characteristics in Recovery Record response clusters.**

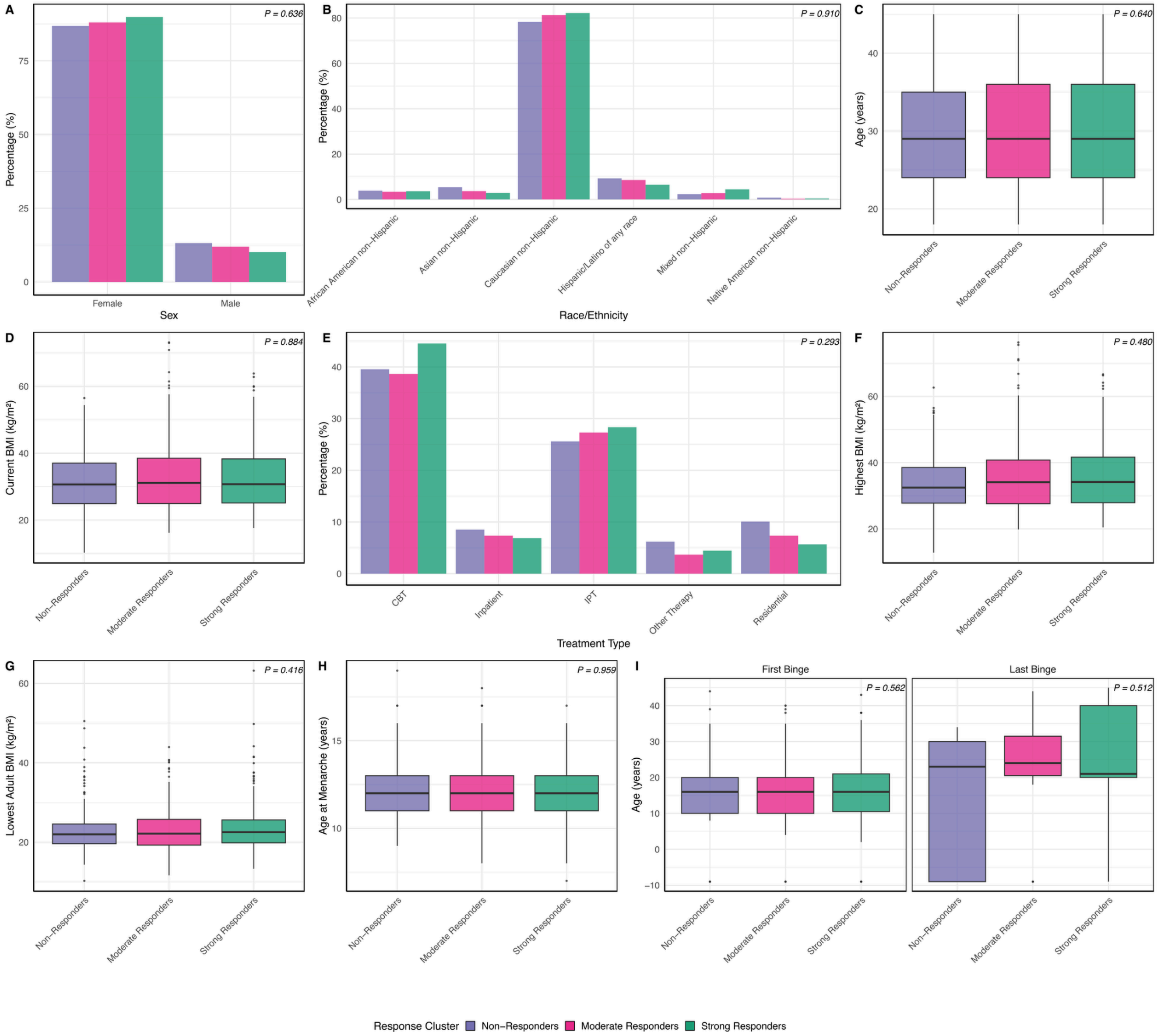


*Ref*=reference, *ns*=not significant, *=*P*<0.05, **=*P*<0.01, ***=*P*<0.001.

*The proportion (or distribution) of individuals among Recovery Record non-responders, moderate responders, and strong responders by sex (A), race (B), age (C), current weight (D), treatment type (E), highest body mass index (BMI) (F), lowest adult BMI (G), age at menarche (among females; H), and age at first and last binge (I). Global P-values are reported for each panel, which were not significantly different between response clusters.*
